# Comparative Performance of agentic AI and Physicians in Taking Clinical History across Leading Large Language Models (LLMs)

**DOI:** 10.64898/2026.01.23.26344723

**Authors:** Sophie Steinbuch, Luka de Vos-Hillebrand, Connor O’Neill-Dee, Iris Wu, Hannah Steinbuch, Zsófi Kulcsár, Avanish Ranjan, Ashish Verma, Jennifer Landsberg, Dimo Dietrich, Charles C. Hardin, Rakesh K. Jain, Sonu Subudhi

**Affiliations:** Steele Laboratories of Tumor Biology, Department of Radiation Oncology, Massachusetts General Hospital and Harvard Medical School, Boston, United States; Clinic for of Dermato-Oncology and Phlebology, University Hospital Bonn, University of Bonn, Bonn, Germany; Clinic and Polyclinic for Ear, Nose and Throat Medicine, University Hospital Bonn, University of Bonn, Bonn, Germany; Department of Medicine (Nephrology), Boston University Chobanian & Avedisian School of Medicine, Boston, United States; Harvard-MIT Health Sciences and Technology Program, Harvard Medical School, Boston, United States; Medicine-Pulmonary and Critical Care Medicine, Massachusetts General Hospital, Boston, United States; Department of Internal Medicine, University Hospital Zürich, Zürich, Switzerland; Adobe, Boston, United States; Koch Institute for Integrative Cancer Research, Massachusetts Institute of Technology, Cambridge, United States; Broad Institute of MIT and Harvard, Cambridge, United States

**Keywords:** Large language models, agentic AI, clinical history-taking, pre-visit workflows, clinical decision support, electronic health records, patient–clinician interaction

## Abstract

Comprehensive clinical history taking is essential for high-quality care. We hypothesized that large language models (LLMs), guided by a structured agentic framework, can efficiently obtain clinically meaningful patient histories. We developed an iterative prompting system that evaluates relevance and completeness across standard history domains and generates targeted follow-up questions until sufficient detail is obtained. We built a patient-facing application and evaluated it using 52 published case reports and 20 constructed clinical scenarios with simulated patient interactions. The framework was implemented using GPT-4o, Gemini-2.5-Flash-Lite, or Grok-3. After each interaction, the system generated an EHR-ready clinical summary, differential diagnosis, and recommended investigations. Across models, relevant history elements were captured with >85% accuracy and F1 scores, as independently assessed by three blinded physicians, and recommended investigations aligned with those used to establish final diagnoses. These findings support the potential of agentic LLM systems for structured clinical history collection and justify prospective clinical evaluation.

## MAIN TEXT

Clinical history is a foundational pillar of diagnostic reasoning, triage and treatment planning. However, outpatient clinicians frequently operate under substantial time constraints and documentation burden, which can lead to incomplete histories, diagnostic delays and fragmented clinical communication. Systems powered by large language models (LLMs) have therefore emerged as promising tools to augment history-taking by improving both efficiency and patient engagement^1,2^.

Recent studies highlight both the strengths and limitations of LLMs in clinical settings. While state-of-the-art models perform well on medical examination questions and synthetic case vignettes, their performance is less consistent in realistic clinical tasks such as collecting comprehensive patient histories^3^. For example, general-purpose LLMs have been shown to underperform resident physicians in emergency department scenarios, often producing conservative or insufficiently precise diagnostic, triage, and management recommendations^4^. By contrast, purpose-built clinical systems that employ structured conversational strategies have demonstrated improved diagnostic dialogue and differential diagnosis generation, in some cases matching or exceeding clinician performance in simulated evaluations^5–7^. These findings suggest that the limitations of LLMs in history-taking may reflect the design of conversational frameworks rather than intrinsic model capability.

As LLM-based systems begin to enter clinical workflows, rigorous methods for evaluating their ability to collect clinically meaningful histories are essential. Prior evaluations indicate that unstructured conversational designs can lead to omissions and variability in collected information^8^. Additionally in clinical settings, expert-led validation remains the gold standard for assessing AI performance, as human oversight provides a level of contextual scrutiny that autonomous self-evaluation currently cannot replicate. Structured, agentic AI systems that explicitly guide dialogue through modular clinical tasks provide a principled approach to both assessment and improvement^9^. Moreover, recent advances allow such systems to be deployed within secure, HIPAA-compliant environments, supporting their potential use in healthcare settings^10,11^.

To enable systematic pre-visit history collection, we developed a conversational application that employs a structured, modular prompting framework. The system progresses through the major components of a clinical history taking process in a predefined sequence, including history of present illness, past medical history, medication history, and social and family history. For each section, the agent generates targeted questions, evaluates the clinical relevance of patient responses, records accepted information, produces interim summaries, and determines whether sufficient detail has been obtained for each section or whether additional follow-up questioning is required (Extended Data Fig. 1). Separate interfaces are provided for patients and physicians. Patients engage with a web-based conversational intake interface that supports iterative, context-aware history collection in real time, whereas clinicians review a structured, EHR-ready summary that consolidates the captured history and presents diagnostic impressions and recommended investigations for downstream clinical assessment (Extended Data Fig. 2). The application is deployed as an online system, with access details provided in the Online Content section.

Using this framework, we benchmarked three leading LLMs (GPT-4o, Grok-3 and Gemini-2.5-Flash-Lite) across fifty-two published clinical case reports drawn from peer-reviewed medical journals, spanning thirteen medical specialties, as well as twenty constructed clinical scenarios designed to represent common outpatient presentations and routine diagnostic decision contexts that are underrepresented in published case reports, which often emphasize rare or atypical conditions (Extended Data Table 1). Simulated patient interactions were conducted by physicians and medical students following the source case descriptions. Independent clinician evaluators from different institutions then assessed the chatbot-generated histories against the corresponding gold-standard case reports using a predefined rubric, with evaluators blinded to model identity.

Across the published case reports, all three models demonstrated consistently high performance in structured history collection. Overall F1 score and recall were high and tightly clustered across models, with narrow confidence intervals, indicating stable and reproducible behavior across cases and evaluators (Fig. 1a). These results show that the structured agentic framework enables reliable capture of clinically relevant history elements and minimizes variability associated with free-form conversational designs. Clinician assessments showed modest but consistent agreement in relative model performance across cases (moderate criterion validity ICCs: 0.50–0.58), despite poor inter-rater reliability among clinicians, likely reflecting the inherent subjectivity of flexible, clinician-defined scoring across history components.

**Figure 1.**
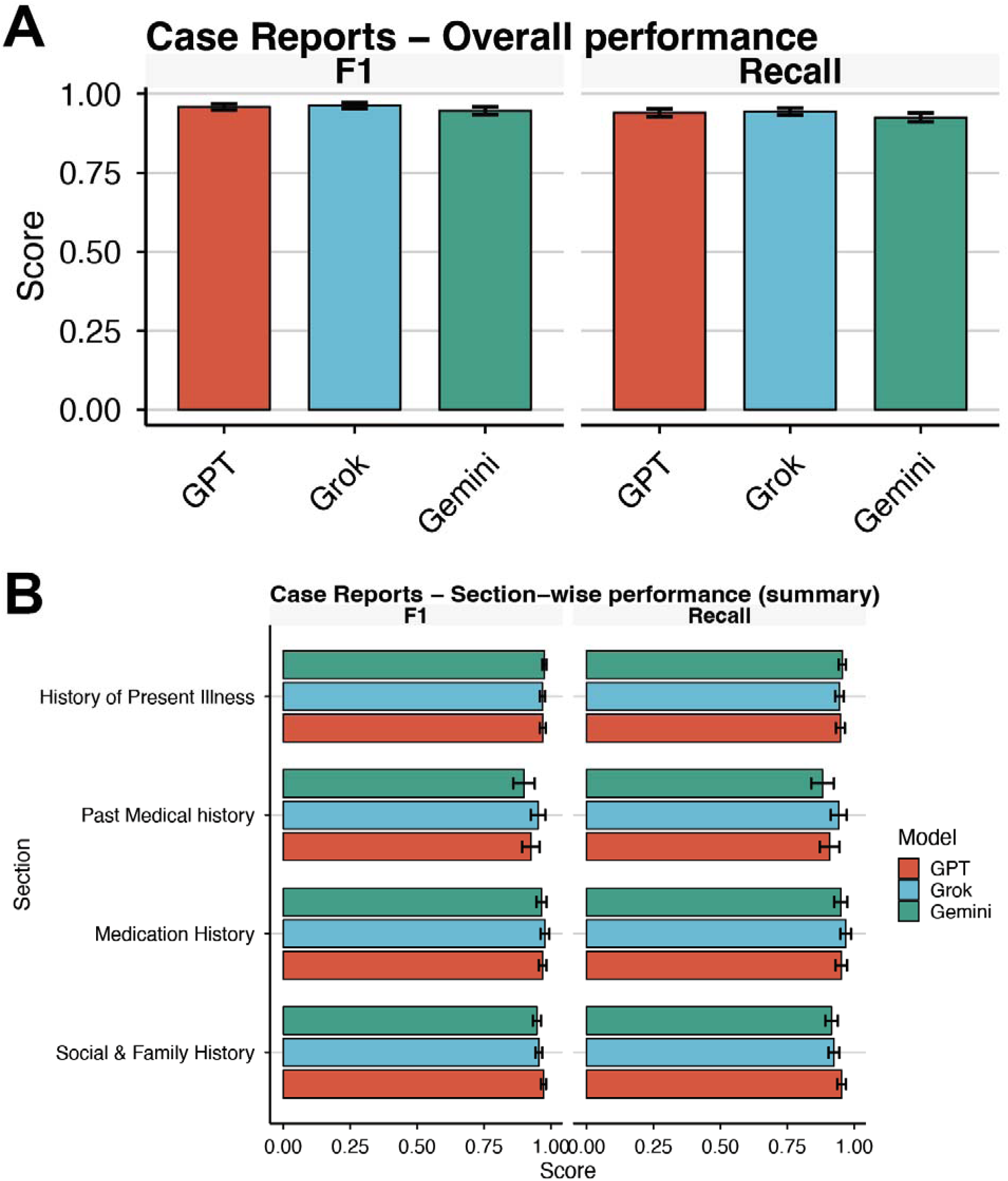
Overall history collection and downstream reasoning performance on published case reports. **(a)** Overall performance of the structured chatbot across published clinical case reports, shown as mean F1 score and recall for three large language models (GPT, Grok and Gemini). Scores are averaged across all evaluated cases and clinician reviewers; error bars denote 95% confidence intervals, indicating stable and reproducible history collection across models. **(b)** Clinician-assessed downstream reasoning performance on published case reports, including generation of a likely diagnosis and identification of clinically important red flags. Bars represent mean normalized scores (0–1) across cases and evaluators, with error bars indicating 95% confidence intervals.

We next examined performance across individual components of the clinical history. All models showed strong and uniform results across history of present illness, past medical history, medication history, and social and family history, with consistently high section-wise F1 score and recall (Fig. 1b). This indicates that the framework promotes balanced coverage of clinical content rather than selective optimization of a subset of history elements. Case-level section-wise analyses further demonstrated that no single history component systematically underperformed across cases or models, supporting the robustness of section-level performance (Extended Data Fig. 3a).

To assess generalizability, we evaluated the same framework on twenty constructed clinical scenarios representing common outpatient presentations with a broader range of presenting complaints, comorbidities, and clinical trajectories. Overall performance on these constructed cases was qualitatively similar to that observed for the published case reports, with high F1 score and recall across all three models and narrow confidence intervals despite increased heterogeneity in case design (Fig. 2a). Section-wise performance across constructed cases remained stable across history components, with variability comparable to that observed in the published case reports (Extended Data Fig. 3b). Together, these findings indicate that the framework generalizes beyond the structure of published case reports and performs robustly across both naturalistic and commonly encountered clinical scenarios.

**Figure 2.**
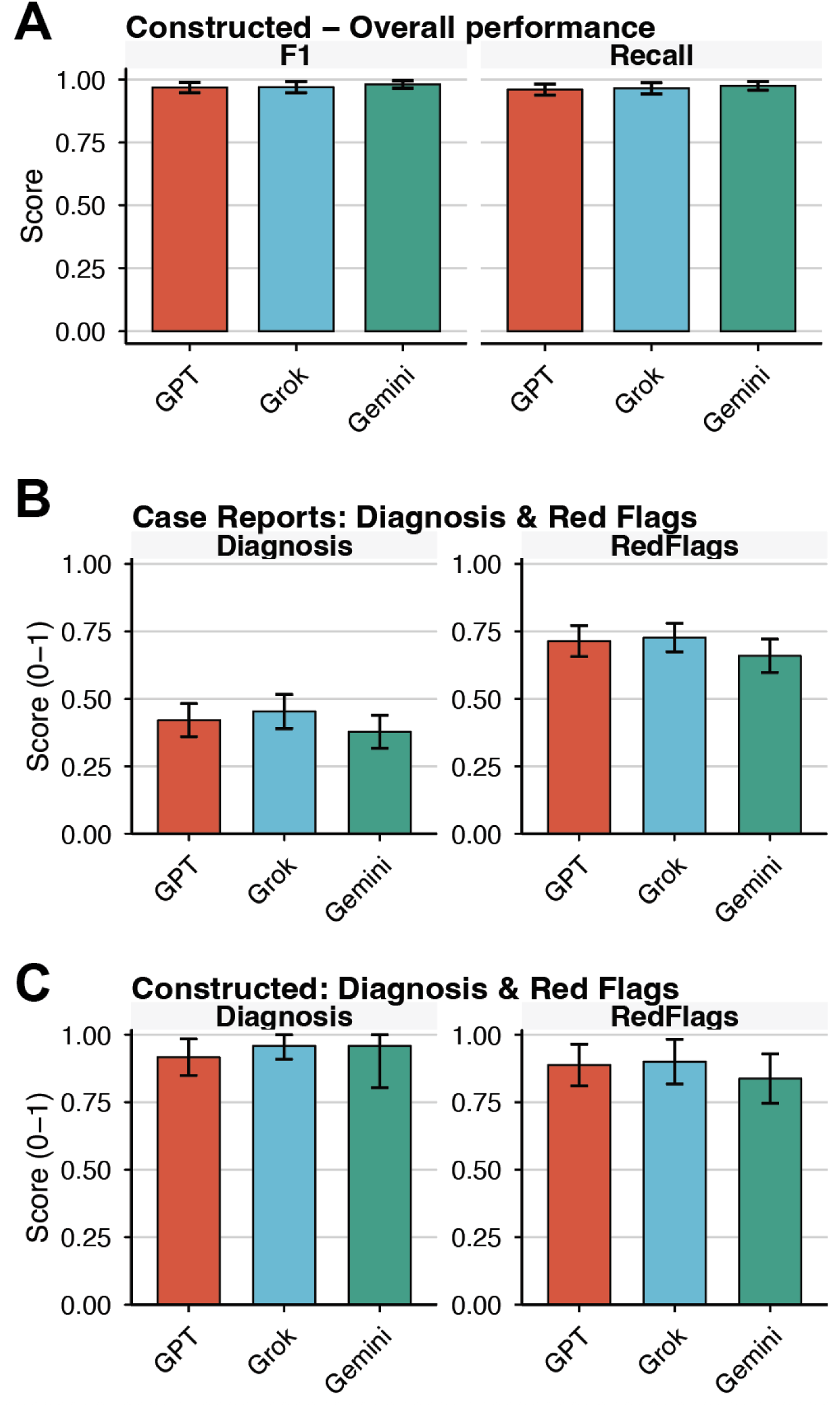
Section-wise robustness and generalizability of structured history collection. **(a)** Section-wise performance on published case reports, stratified by major components of the clinical history (history of present illness, past medical history, medication history, and social and family history). Mean F1 score and recall are shown for each model; error bars represent 95% confidence intervals. Metrics are computed conditional on sections containing clinically relevant information, demonstrating balanced coverage across history domains. **(b)** Overall performance on constructed clinical scenarios representing common outpatient presentations, shown as mean F1 score and recall across all sections and clinician evaluators, with 95% confidence intervals. **(c)** Clinician-assessed diagnostic performance and identification of clinically important red flags for constructed cases. Bars show mean normalized scores (0–1) across cases and evaluators, with error bars indicating 95% confidence intervals.

We next evaluated downstream clinical reasoning tasks, specifically generation of a likely diagnosis and identification of clinically important red flags. Across the published case reports, diagnostic performance was moderate for all three models and consistently lower than history collection metrics (Fig. 2b). This finding is expected, as accurate diagnosis often requires integration of physical examination findings, laboratory data, imaging, and disease progression, which were not available to the LLMs. By contrast, performance on red-flag identification was substantially stronger, with all models reliably highlighting major safety concerns (Fig. 2b).

Evaluation of the constructed cases yielded a distinct pattern. Diagnostic performance was higher for these scenarios, which were intentionally designed to reflect common clinical presentations that are relatively straightforward to diagnose (Fig. 2c). Identification of red flags remained consistent and within an acceptable range across models, even in cases with overlapping symptom patterns or diagnostic ambiguity (Fig. 2c).

Finally, pairwise comparison of aggregate history scores revealed broadly concordant performance across models for both published case reports and constructed clinical scenarios (Extended Data Fig. 4). Across the published case reports, Grok demonstrated modest but statistically significant differences relative to GPT and Gemini, whereas no significant difference was observed between Gemini and GPT. In the constructed cases, pairwise differences were smaller and did not consistently reach statistical significance, reflecting increased overlap in model performance across common clinical scenarios. These analyses indicate that while small model-specific differences exist, total history capture is largely comparable across models when deployed within the structured agentic framework.

Taken together, these findings demonstrate that a structured agentic framework substantially improves the ability of large language models to collect clinically meaningful patient histories across a wide range of scenarios. Across seventy-two total clinical cases, encompassing both published case reports and constructed clinical scenarios, the system achieved high and stable performance across all major components of the clinical history and reliably identified dangerous conditions requiring consideration. Although diagnostic performance was more heterogeneous as expected when relying on history alone, the framework consistently supported clinically relevant safety assessment.

These results have important implications for clinical practice. Automated, structured history collection has the potential to improve completeness of patient information, decrease omissions that contribute to delayed or missed diagnoses and most importantly reduce documentation burden, thereby reducing physician burnout. The observed stability of performance across diverse cases suggests that such systems could function as reliable intake tools in outpatient clinics, telemedicine encounters, and urgent care settings. The modular design of the agentic framework further enables adaptation to specialty-specific workflows and integration with electronic health record systems.

This study has several limitations. All encounters were simulated rather than derived from real patient interactions, which may not fully capture the variability of communication observed in clinical practice. In addition, clinician reviewers noted that in a subset of cases, the system did not consistently elicit sex-specific or context-dependent history elements; for example, pregnancy-related questions in young women presenting with abdominal pain. However, this reflects limitations of the current prompting strategy rather than fundamental model capability and could be addressed in future iterations through targeted prompt constraints, rule-based safety checks, or section-specific branching logic.

Although more advanced large language models were available at the time of evaluation, we prioritized models with lower inference latency to support responsive, iterative dialogue; more computationally intensive models may improve downstream reasoning but can introduce delays that are impractical for real-time clinical intake. Finally, the scale of evaluation was constrained by the use of manual clinician review for each generated history. This rigorous assessment approach limited the total number of cases relative to studies that rely on automated self-evaluation by LLMs but ensured clinically meaningful and independent validation of performance. Future studies should evaluate the framework in real-world clinical deployments, incorporate multimodal clinical data such as laboratory results and imaging, and assess generalizability across a broader range of model architectures.

## Data Availability

The datasets generated and analyzed during this study include published clinical case reports and synthesized clinical scenarios. Published case reports are publicly available from their original sources. Synthesized clinical scenarios were constructed by the authors to represent common outpatient presentations and do not contain real patient data; details of cases are provided in Extended Data Table 1.
The core conversational agent software used for clinical history collection is subject to a pending patent application and is therefore not publicly available at this time. Code used for data processing, metric computation, statistical analysis, and figure generation will be made publicly available upon publication via a version-controlled GitHub repository.

https://github.com/sonusubudhi/agentic_AI_chatbot_R_code

## METHODS

### Study Design and Overview

We designed a multi-stage evaluation to assess whether a structured agentic artificial intelligence (AI) system can reliably collect pre-visit clinical histories using large language models (LLMs). Three LLMs were evaluated using this agentic framework: GPT-4o, Grok-3, and Gemini-2.5-flash-lite. The study consisted of two components. First, applying each LLM, we evaluated the system on fifty-two published clinical case reports drawn from peer-reviewed journals spanning thirteen medical specialties. Similarly, we evaluated all three LLMs on twenty constructed clinical scenarios designed to represent common outpatient presentations and to complement the breadth of the published cases (Extended Data Table 1). All cases were evaluated using simulated patient–chatbot interactions conducted by physicians and medical students following the source case descriptions.

### Description of the Agentic AI System

The agentic AI system was designed to guide LLMs through a modular, clinically grounded workflow for structured history collection. The system comprises discrete modules for question generation, response validation, short-term and long-term memory management, interim summarization, follow-up question selection, and conversation termination. These modules communicate through explicit internal instructions, allowing fine-grained control over the interaction flow. The system progresses sequentially through the major components of a standard clinical history, including the chief complaint, history of present illness, past medical history, medication history, allergies, social and family history, and review of systems. Within each section, patient responses are evaluated for clinical relevance and completeness before being stored in a structured history state. Interim summaries are generated to maintain coherence and contextual awareness. At the conclusion of each interaction, the system produces an electronic health record (EHR)-compatible clinical summary, a list of likely diagnoses, a list of dangerous diagnoses requiring exclusion, and recommended initial investigations. The agentic control logic was held constant across all evaluations, with only the underlying LLM varied between experiments (Extended Data Fig. 1).

### Simulated Patient Interactions

Simulated patient interactions were conducted by physicians and advanced medical students who assumed the role of the patient while referencing the full content of each case. Participants were instructed to respond conversationally, provide only information contained in the source case, and avoid introducing new details. Each case was evaluated independently with each model. This resulted in 156 total chatbot-generated history transcripts for the 52 published case reports and 60 transcripts for the twenty constructed cases.

### Reference Standards

For published case reports, the gold-standard history and final diagnosis were derived directly from the original publication. For constructed cases, the gold-standard history and diagnosis corresponded to the scenario intentionally designed by the research team. Clinically important red flags were defined as diagnoses or conditions requiring urgent exclusion or immediate clinical action, based on consensus among clinician reviewers and the safety considerations described in each case.

### Clinician Scoring Procedure

Three trained physicians independently evaluated each chatbot-generated history alongside the corresponding gold-standard history. Scoring was performed using a predefined rubric that assessed the presence and clinical relevance of key history elements within each section of the encounter, including history of present illness, past medical history, medication history, and social and family history (Extended Data Table 2). Metrics were computed conditional on sections containing clinically relevant information in the reference case. Reviewers also evaluated system-generated diagnostic suggestions and identification of clinically important red flags relative to the reference answers. All scores were aggregated across reviewers for downstream analysis.

### Evaluation Metrics

Performance was primarily assessed using recall and F1 score, which quantify the completeness and balance of captured clinical information. Metrics were computed for the full clinical history and for individual history sections. For diagnostic performance and red-flag identification, normalized scores ranging from 0 to 1 were computed based on clinician grading. Summary statistics are reported as mean values with 95% confidence intervals across cases and evaluators.

## Statistical Analysis

Scores were aggregated across clinician evaluators for each case prior to analysis. Primary analyses focused on relative model performance rather than absolute score calibration. Between-model comparisons were primarily descriptive. Exploratory pairwise comparisons of aggregate history scores between models were performed using paired Wilcoxon signed-rank tests, with corresponding *P* values reported where applicable (Extended Data Fig. 4).

To assess inter-rater reliability and criterion validity, clinician agreement was quantified using the intraclass correlation coefficient (ICC). Because clinicians applied individualized scoring scales and weightings across history components, raw scores were first Z-score normalized within each clinician to enable meaningful comparison of relative rankings. Inter-rater reliability was evaluated separately for the original NEJM histories and for each AI model using a two-way random-effects ICC for consistency, averaged across raters (ICC(C,k)). Criterion validity was assessed by comparing each AI model’s normalized scores to the normalized original history scores using a two-way random-effects ICC for consistency, single rater (ICC(C,1)). This approach captures consistency in relative performance while fully accommodating differences in absolute scoring scales and clinician preferences. All statistical analyses and figure generation were performed using R (version 4.5.1).

## Ethical Considerations

This study used only published clinical case reports and synthesized scenarios and did not involve real patient data. As such, it did not constitute human subjects research under United States federal regulations and did not require institutional review board approval.

## DATA AND CODE AVAILABILITY

The datasets generated and analyzed during this study include published clinical case reports and synthesized clinical scenarios. Published case reports are publicly available from their original sources. Synthesized clinical scenarios were constructed by the authors to represent common outpatient presentations and do not contain real patient data; details of cases are provided in Extended Data Table 1.

The core conversational agent software used for clinical history collection is subject to a pending patent application and is therefore not publicly available at this time. Code used for data processing, metric computation, statistical analysis, and figure generation will be made publicly available upon publication via a version-controlled GitHub repository.

## ONLINE CONTENT

Access to the web-based structured history-taking application is available at https://clinicalchatbot-hdhceeejc4ahe6cv.eastus2-01.azurewebsites.net. The system was used solely for research evaluation.

## ACKNOWLEDGEMENTS

We thank the members of the Edwin L. Steele laboratories for helpful discussions. S. Subudhi was supported by the MGH ECoR FMD fellowship grant (2022A018897). R.K.J. was supported by grants from the NIH (R13CA306205, R01CA269672, U01CA261842 R01CA259253, and U01CA224348), the Ludwig Cancer Center at Harvard, the Nile Albright Research Foundation, the National Foundation for Cancer Research and the Jane’s Trust Foundation.

## Extended Data Figures

**Extended Data Figure 1.**
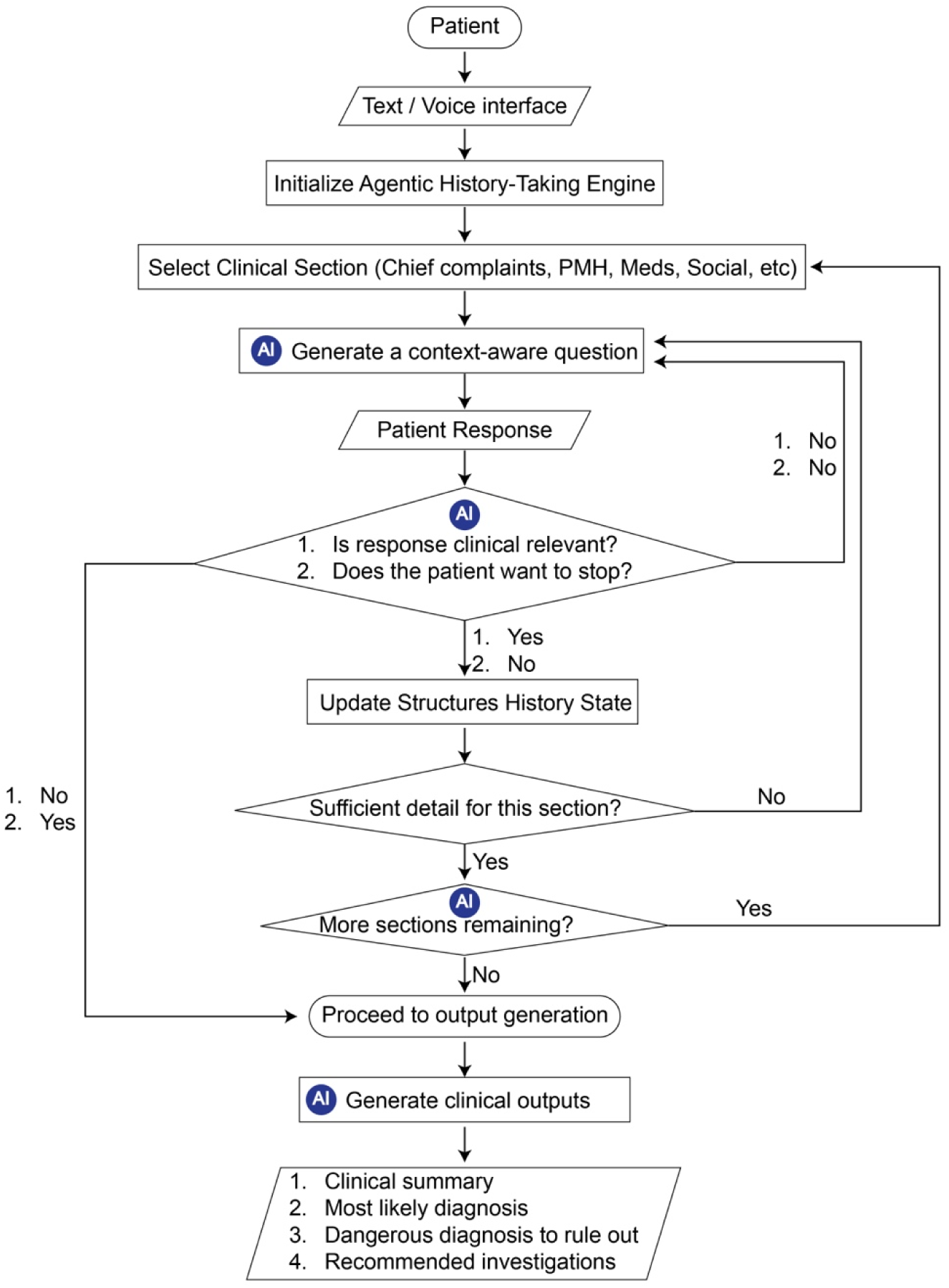
Agentic framework for structured clinical history collection. Schematic overview of the agentic history-taking engine. Following patient initiation via a text or voice interface, the system sequentially selects clinical history sections (for example, chief complaint, history of present illness, past medical history, medication history, and social and family history). For each section, the agent generates context-aware questions, evaluates the clinical relevance of patient responses, updates a structured history state, and determines whether sufficient detail has been obtained or whether additional follow-up is required. Once all sections are completed or the encounter is terminated, the system generates structured clinical outputs, including a clinical summary, likely diagnosis, dangerous diagnoses to rule out, and recommended investigations.

**Extended Data Figure 2.**
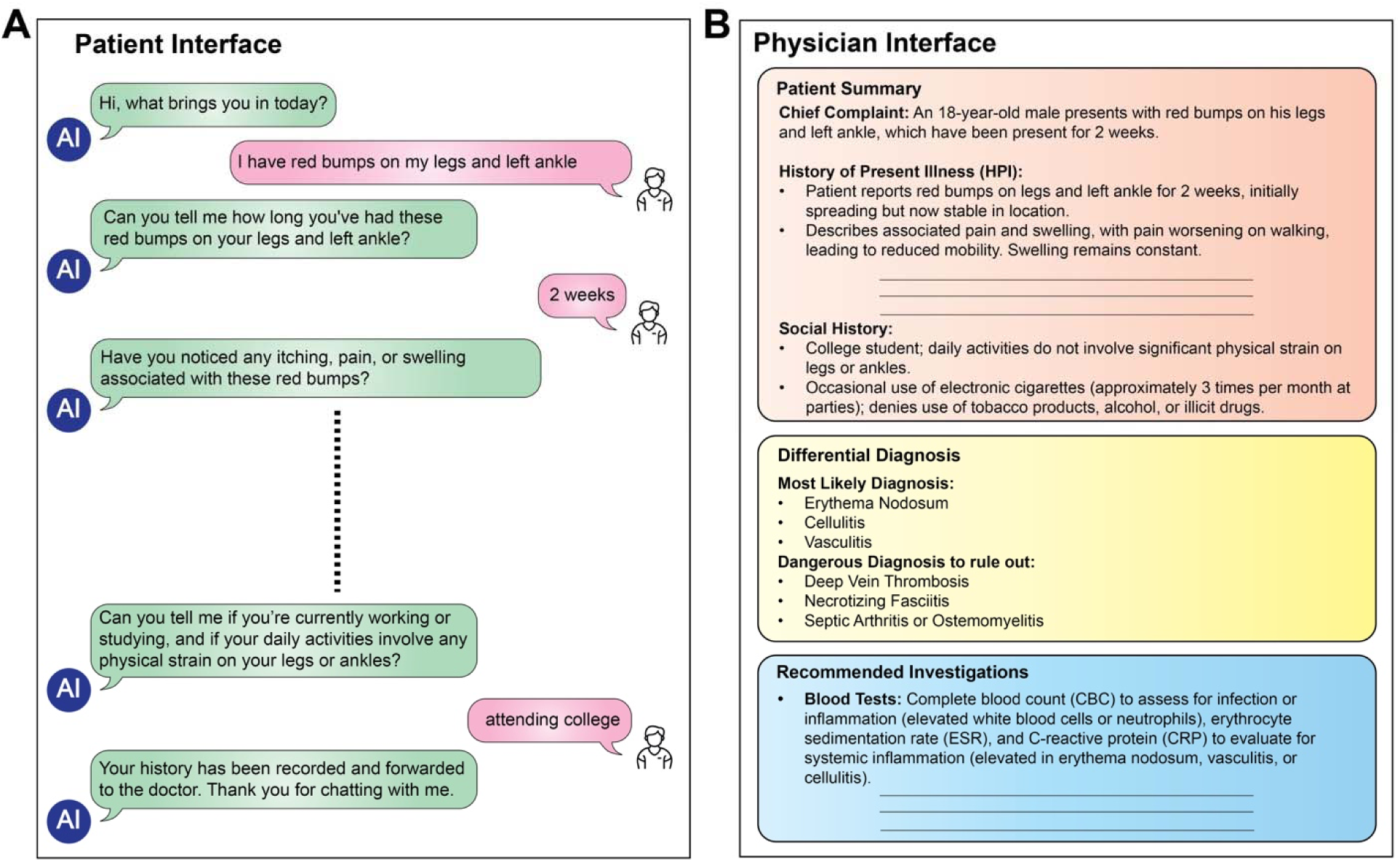
Patient-facing and physician-facing interfaces of the structured chatbot. **(a)** Patient-facing conversational interface illustrating iterative, context-aware questioning during history collection. **(b)** Physician-facing interface displaying the structured clinical summary generated by the system, differential diagnosis, dangerous diagnoses to rule out, and recommended initial investigations. Outputs are formatted to be compatible with downstream clinical review and documentation.

**Extended Data Figure 3.**
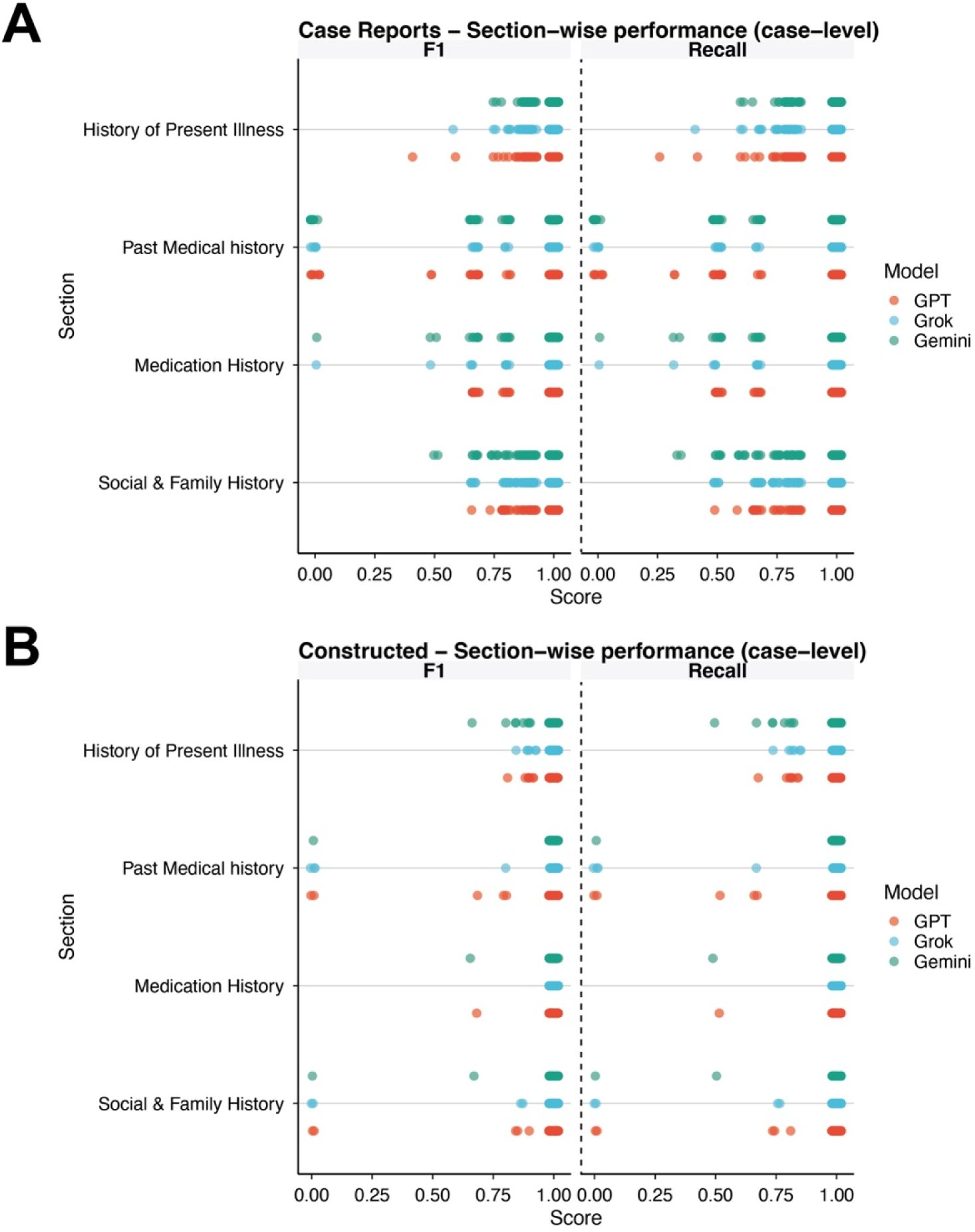
Case-level section-wise performance across clinical history components. Case-level distribution of F1 score and recall across individual history sections for published case reports (a) and constructed clinical cases (b). Points represent individual case evaluations for each model, stratified by history of present illness, past medical history, medication history, and social and family history. These plots illustrate variability across cases and sections, complementing the summary statistics shown in the main figures.

**Extended Data Figure 4.**
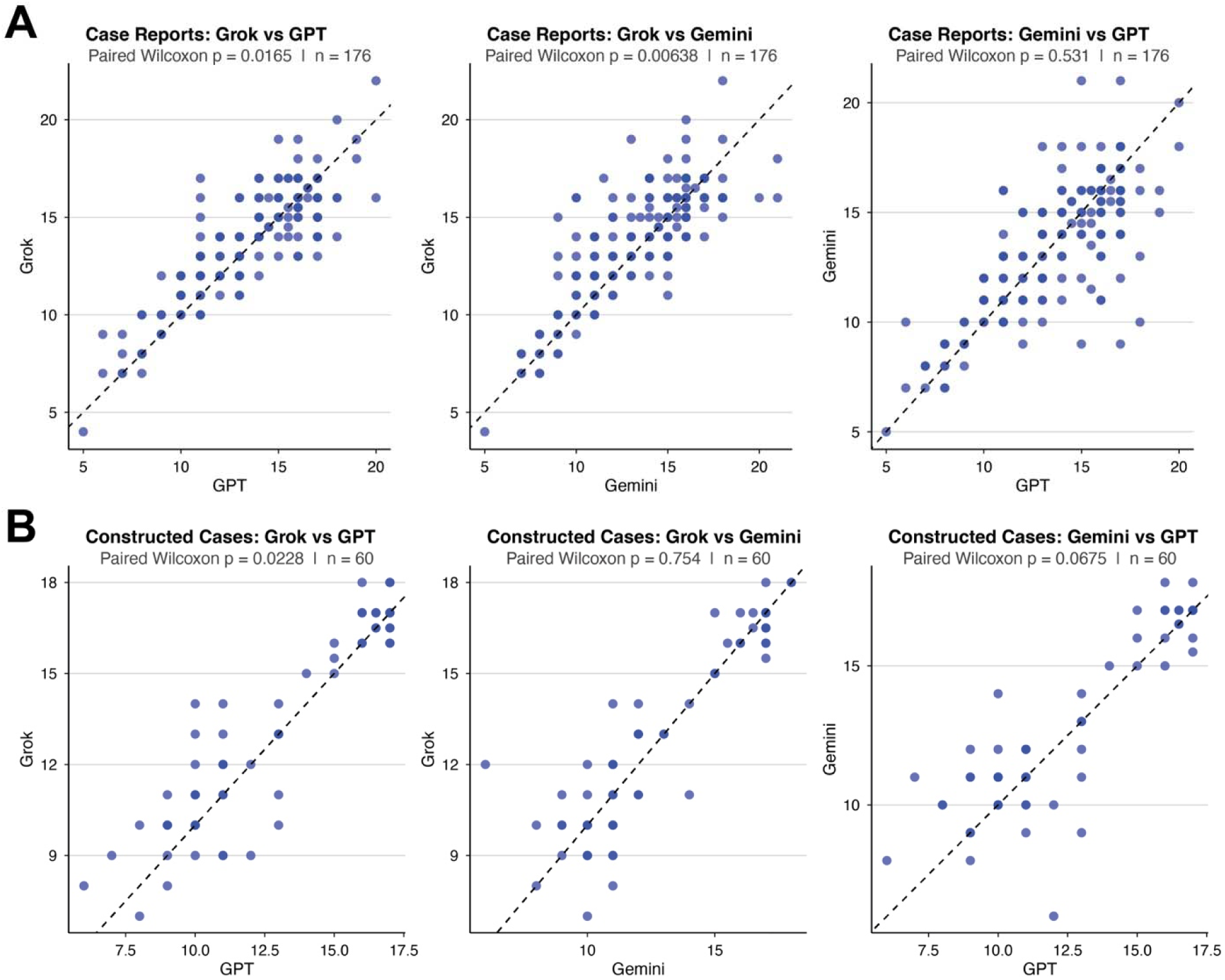
Pairwise comparison of raw history scores between large language models. Pairwise scatter plots comparing total history scores assigned to chatbot-generated histories across models for published case reports (a) and constructed clinical cases (b). Each point represents a single case evaluated by a clinician, plotted against the identity line (dashed). Paired Wilcoxon signed-rank tests were used to assess differences between model pairs, with corresponding P values and sample sizes indicated. Scores reflect aggregate clinician grading across all evaluated history components.

